# Surveillance of COVID-19 vaccine safety among elderly persons aged 65 years and older

**DOI:** 10.1101/2022.11.04.22281910

**Authors:** Hui-Lee Wong, Ellen Tworkoski, Cindy Ke Zhou, Mao Hu, Deborah Thompson, Bradley Lufkin, Rose Do, Laurie Feinberg, Yoganand Chillarige, Rositsa Dimova, Patricia Lloyd, Thomas MaCurdy, Richard A. Forshee, Jeffrey Kelman, Azadeh Shoaibi, Steven A. Anderson

**Author notes:** **Corresponding Author:** Steven A. Anderson, PhD, (SA).

## Abstract

**Background:** Monitoring safety outcomes following COVID-19 vaccination is critical for understanding vaccine safety especially when used in key populations such as elderly persons age 65 years and older who can benefit greatly from vaccination. We present new findings from a nationally representative early warning system that may expand the safety knowledge base to further public trust and inform decision making on vaccine safety by government agencies, healthcare providers, interested stakeholders, and the public.

**Methods:** We evaluated 14 outcomes of interest following COVID-19 vaccination using the US Centers for Medicare & Medicaid Services (CMS) data covering 30,712,101 elderly persons. The CMS data from December 11, 2020 through Jan 15, 2022 included 17,411,342 COVID-19 vaccinees who received a total of 34,639,937 doses. We conducted weekly sequential testing and generated rate ratios (RR) of observed outcome rates compared to historical (or expected) rates prior to COVID-19 vaccination.

**Findings:** Four outcomes met the threshold for a statistical signal following Pfizer-BioNTech vaccination including pulmonary embolism (PE; RR=1.54), acute myocardial infarction (AMI; RR=1.42), disseminated intravascular coagulation (DIC; RR=1.91), and immune thrombocytopenia (ITP; RR=1.44). After further evaluation, only the RR for PE still met the statistical threshold for a signal; however, the RRs for AMI, DIC, and ITP no longer did. No statistical signals were identified following vaccination with either the Moderna or Janssen vaccines.

**Interpretation:** This early warning system is the first to identify temporal associations for PE, AMI, DIC, and ITP following Pfizer-BioNTech vaccination in the elderly. Because an early warning system does not prove that the vaccines cause these outcomes, more robust epidemiologic studies with adjustment for confounding factors, including age and nursing home residency, are underway to further evaluate these signals. FDA strongly believes the potential benefits of COVID-19 vaccination outweigh the potential risks of COVID-19 infection.

## Introduction

The US Food and Drug Administration (FDA) is monitoring the safety of three vaccines for Coronavirus Disease 2019 (COVID-19) currently available in the US. These include the licensed Pfizer BioNTech vaccine (Comirnaty) for persons 16 years and older and authorized under emergency use authorization (EUA) for those 12-15 years (BNT162b2), as well as the authorized Moderna (mRNA-1273), and Janssen (Ad26.COV2.S) vaccines for persons 18 years and older. Pre-authorization clinical studies provided useful information on the safety of COVID-19 vaccines, but they have limitations such as sample size and follow-up that may be addressed in post-authorization safety studies in large healthcare databases. Accordingly, FDA and the Centers for Medicare & Medicaid Services (CMS) are using the Medicare health insurance database, covering more than 25 million elderly persons aged 65 years and older, to conduct near real-time safety monitoring of 14 outcomes on a weekly basis following COVID-19 vaccine administration.

The US elderly population, including persons in Long-Term Care Facilities or nursing homes [1], were disproportionately affected by COVID-19, as they were among the first US populations to be infected. Because they suffered a higher rate of infection, serious disease, and severe outcomes including death [2], they were among the first groups recommended by the Advisory Committee on Immunization Practices to receive the vaccine [3]. Available information concerning the safety of the vaccine in elderly persons is limited. However, the near real-time surveillance method used by FDA and CMS continues to expand the available knowledge base and further advances our understanding of the safety profile of these new COVID-19 vaccines. The FDA-CMS near real-time active surveillance program complements other FDA and US Government vaccine safety surveillance systems by rapidly detecting safety concerns that may not have been voluntarily reported to passive surveillance systems such as Vaccine Adverse Event Reporting System. Routinely used by FDA in safety surveillance for annual influenza vaccines in the past decade, this method is designed to be sensitive enough to quickly screen safety signals for further evaluation in robust epidemiologic studies [4, 5]. This rapid screening method performs hypothesis testing, sequentially, in a prospective manner as the vaccine data accrue to detect potential safety signals earlier in the course of surveillance, but signals must be further evaluated in more robust studies with adjustment for confounding. However, results detected by near real-time surveillance do not establish a causal association between the outcomes and vaccination because the method has limited adjustments for confounding.

In this report, we summarize the results of weekly sequential testing analyses for 14 outcomes where formal testing was initiated. We also describe how the identified signals were evaluated and how the epidemiological studies currently underway will provide more robust adjustment for confounding determine if any are true signals.

## Methods

### Data Sources

We used the US Medicare Fee-for-Service (FFS) Parts A (inpatient services) and B (outpatient care) claims and enrollment data to define inclusion criteria, exposures, outcomes, and patient characteristics. The number of 65+ beneficiaries with at least 1 day of Medicare FFS enrollment in the study period is 30,712,101. The Minimum Data Set identified nursing home residency status.

### Study Period and Population

The study included Medicare FFS beneficiaries aged 65 years or older who received a COVID-19 vaccine since December 11, 2020. To be included, individuals needed to be enrolled on the vaccination date, and continuously enrolled during an outcome-specific pre-vaccination clean window [6].

### Exposure and Follow-Up

Exposure was defined as receipt of a Pfizer-BioNTech, Moderna, or Janssen COVID-19 vaccination, identified using brand and dose-specific Current Procedural Terminology / Healthcare Common Procedure Coding System codes [7] (Table S1). The primary analysis included all observed doses by brand. Dose-specific analyses are described in supplemental materials. Follow-up included all time in the prespecified risk windows; the first-dose risk window was censored at the time of the second-dose vaccination.

### Outcomes

The list of 14 outcomes, pre-vaccination clean windows, and post-vaccination risk windows are detailed in Table S2. Claims-based outcome algorithms were developed based on literature review and in consultation with clinical experts [8].

### Vaccine Safety Surveillance

Weekly vaccine uptake was monitored by brand. The near real-time surveillance compared the observed number of each outcome in the COVID-19 vaccinated population to an expected number based on the background rate of the outcome in a similar COVID-19 unvaccinated population prior to the pandemic, adjusted for the delay in claims processing and standardized by nursing home residency status, age, sex, and race. We calculated annual background rates within the strata of the standardized variables, where possible, during 2017-2019 (pre-COVID-19) and peri-COVID-19 (January 1, 2020–December 10, 2020) [9] (Table S3). If annual rates in the historical period differed substantially from each other, we selected the minimum rate as a more conservative approach. Otherwise, the median annual rate was selected.

### Statistical Analysis

Poisson Maximized Sequential Probability Ratio Test (PMaxSPRT) was used to detect increased outcome risk following vaccination compared to a historical baseline for 14 outcomes [10-12] (Table S3). Weekly sequential testing for each outcome commenced when a minimum of three cases accrued. One-tailed tests were used, with a null hypothesis that the observed rate was no greater than the historical comparator beyond a prespecified test margin with an overall alpha of 1%. The test margin was selected for each outcome based on expert guidance to avoid minimal risk increases that were unlikely to be clinically relevant. The alpha level was selected to address a large number of tests. A statistical signal occurred if the log likelihood ratio exceeded the critical value, or a threshold for determining whether the result was likely to occur due to chance. Additional details are provided in Table S3 and the study protocol [6].

#### Signal Evaluation

Prespecified signal evaluation analyses were conducted after a statistical signal was observed. Data quality was checked to rule out database errors or changes in event observation as potential sources of the signal. Sensitivity analyses evaluated whether the increase in risk was observed with alternate expected rates in sequential testing – (i) rates from calendar months in the historical period corresponding to those in the study period to address monthly variations in the background rates, and (ii) rates among a subset of Medicare beneficiaries with influenza vaccination in the past year to assess differential healthcare utilization. For pulmonary embolism (PE), additional ad hoc analyses were conducted with (i) the outcome limited to the inpatient setting and (ii) rates from November 1–December 11, 2020 to address changes in rates in the peri-COVID period as alternate expected rates.

Signal characterization assessed the cases and the potential elevated risk. Analyses included (i) identifying clusters in the risk window following dose 1 through temporal scans, (ii) estimating relative risks within demographic strata, (iii) assessing distribution of care settings and diagnosis codes in observation and historical periods, (iv) comparing patient characteristics among vaccinated and overall elderly populations, and (v) reviewing patterns of reimbursement codes by clinicians on a random sample of up to 100 cases per outcome to contextualize the medical histories of patients.

#### Statistical Software

All analyses were conducted using R 4.0.3 (R Foundation for Statistical Computing, Vienna, Austria), SAS v. 9.4 (SAS Institute Inc., Cary, NC, United States), and SaTScan v9.6 (Martin Kulldorff, Boston, MA, United States).

#### Ethical Considerations

This surveillance activity was conducted as part of the FDA public health surveillance mandate.

## Results

### Descriptive Statistics of the COVID-19 Vaccinated Population

From December 11, 2020 through January 15, 2022, 17,088,796 Pfizer-BioNTech, 16,898,376 Moderna, and 634,019 Janssen COVID-19 vaccine doses were found among 30,712,101 eligible individuals in the study period (34,621,191 total vaccine doses including vaccination days with multiple products). The population of Pfizer-BioNTech vaccinees had a slightly higher proportion of older individuals (85 years and older), nursing home residents, and individuals residing in urban areas compared to the general elderly Medicare population (Table 1; Figure 1). Moderna vaccinees exhibited similar characteristics to the general Medicare population, while the population receiving the Janssen vaccine was younger and had fewer nursing home residents.

**Table 1:** Characteristics of Pfizer-BioNTech, Moderna, and Janssen Vaccine Doses Administered among Adults Aged 65 years and Older in Medicare^a^, Dec 11, 2020 to January 15, 2022.

| Patient Characteristic | General 65+ FFS<sup>b</sup> | Moderna<sup>c</sup> | Pfizer | Janssen | SMD - Comparison with General FFS<sup>c</sup> |  |  |
| --- | --- | --- | --- | --- | --- | --- | --- |
|  |  |  |  |  | Moderna | Pfizer | Janssen |
| **Total** | **25,390,578** | **15,761,718** | **15,896,042** | **576,698** |  |  |  |
| **Nursing Home Residency Status** |  |  |  |  |  |  |  |
| Nursing Home Resident | 578,908 (2.3) | 324,991 (2.06) | 711,437 (4.48) | 12,948 (2.25) | 0.02 | **0.12** | 0.00 |
| Non-Nursing Home Resident | 24,811,670 (97.7) | 15,436,727 (97.94) | 15,184,605 (95.52) | 563,750 (97.75) | 0.02 | **0.12** | 0.00 |
| **Age (years)** |  |  |  |  |  |  |  |
| 65-74 | 13,661,915 (53.8) | 8,333,648 (52.87) | 8,080,719 (50.83) | 347,687 (60.29) | 0.02 | 0.06 | **0.13** |
| 75-84 | 8,288,448 (32.6) | 5,369,646 (34.07) | 5,383,100 (33.86) | 166,011 (28.79) | 0.03 | 0.03 | 0.08 |
| 85+ | 3,440,215 (13.5) | 2,058,424 (13.06) | 2,432,223 (15.30) | 63,000 (10.92) | 0.01 | 0.05 | 0.08 |
| **Sex** |  |  |  |  |  |  |  |
| Female | 14,191,641 (55.9) | 9,010,306 (57.17) | 9,324,988 (58.66) | 319,812 (55.46) | 0.03 | 0.06 | 0.01 |
| Male | 11,198,935 (44.1) | 6,751,412 (42.83) | 6,571,054 (41.34) | 256,886 (44.54) | 0.03 | 0.06 | 0.01 |
| **Race/Ethnicity** |  |  |  |  |  |  |  |
| Asian | 529,362 (2.1) | 320,757 (2.04) | 341,530 (2.15) | 10,113 (1.75) | 0.00 | 0.00 | 0.03 |
| Black | 1,728,808 (6.8) | 764,033 (4.85) | 898,467 (5.65) | 33,219 (5.76) | 0.08 | 0.05 | 0.04 |
| Hispanic | 398,426 (1.6) | 146,371 (0.93) | 162,556 (1.02) | 6,578 (1.14) | 0.06 | 0.05 | 0.04 |
| Alaskan Native/Native American | 117,539 (0.5) | 64,557 (0.41) | 67,878 (0.43) | 1,421 (0.25) | 0.01 | 0.01 | 0.04 |
| White | 21,584,886 (85.0) | 13,776,628 (87.41) | 13,714,047 (86.27) | 504,406 (87.46) | 0.07 | 0.04 | 0.07 |
| Other | 448,630 (1.8) | 282,212 (1.79) | 299,951 (1.89) | 7,555 (1.31) | 0.00 | 0.01 | 0.04 |
| Missing/Unknown | 582,927 (2.3) | 407,160 (2.58) | 411,613 (2.59) | 13,406 (2.32) | 0.02 | 0.02 | 0.00 |
| **Urban/Rural** |  |  |  |  |  |  |  |
| Urban | 19,415,683 (76.5) | 11,855,600 (75.22) | 1,346,1164 (84.68) | 436,146 (75.63) | 0.03 | **0.21** | 0.02 |
| Rural | 5,811,513 (22.9) | 3,822,143 (24.25) | 2,359,912 (14.85) | 138,341 (23.99) | 0.03 | **0.21** | 0.03 |
| Missing/Unknown | 163,382 (0.6) | 83,975 (0.53) | 74,966 (0.47) | 2,211 (0.38) | 0.01 | 0.02 | 0.03 |

Abbreviations: FFS, Fee-for-Service; SMD, standardized mean difference; AMI, acute myocardial infarction; COPD, chronic obstructive pulmonary disease; COVID-19, coronavirus disease 2019
<sup>a</sup> Individuals included in this table were required to have 365 days of continuous enrollment prior to vaccination date in order to accurately capture medical history. Therefore, the total vaccine doses for this subpopulation is smaller than the study population of Medicare FFS 65+ study population reported in the Results section.
<sup>b</sup> Characteristics of the general Medicare FFS population aged 65 years and older were assessed as of 12/10/2020
<sup>c</sup> SMD with values greater than 0.1 are presented in bold text and indicate covariates with larger imbalances between populations

**Fig 1.**
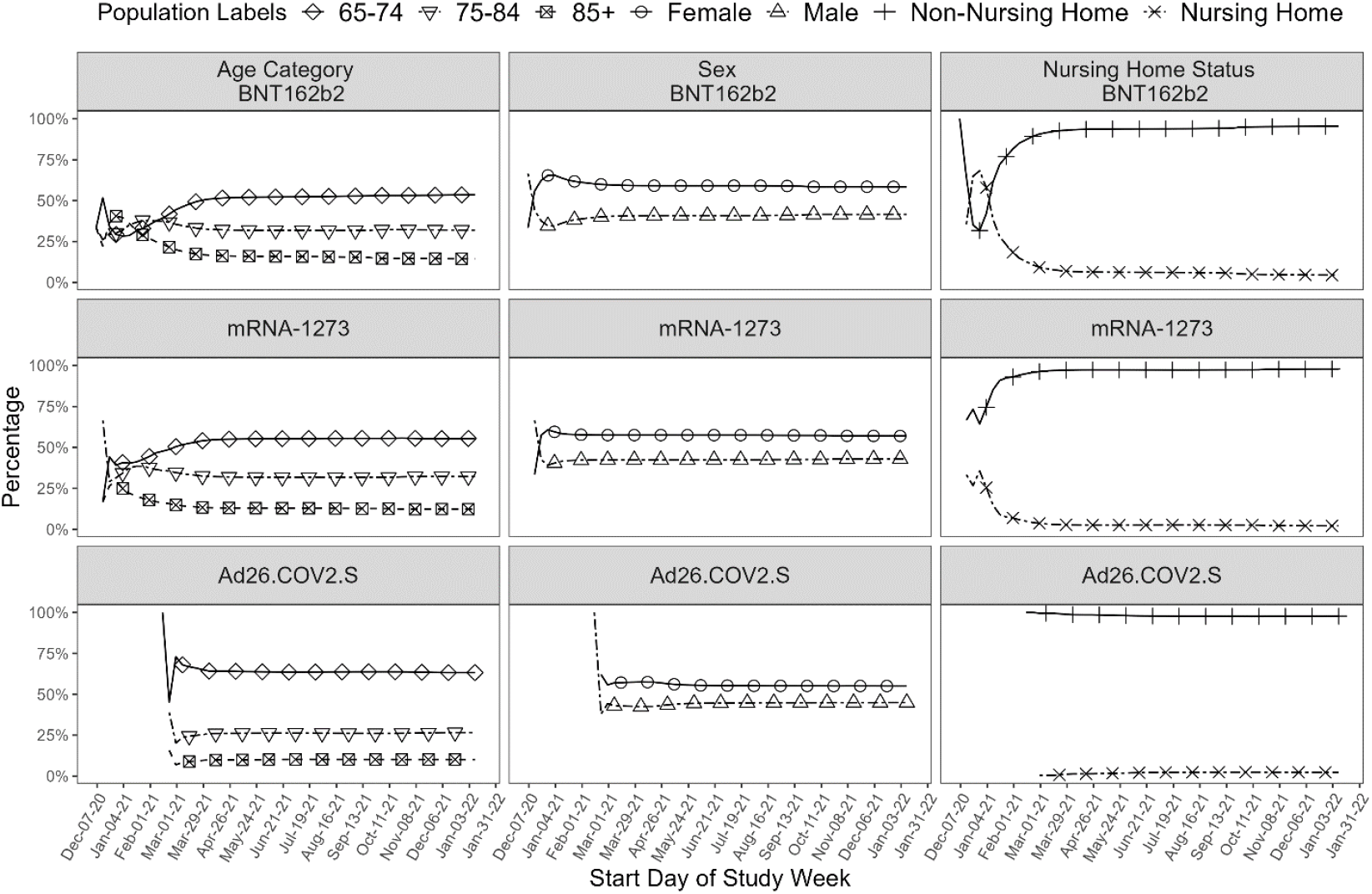
Cumulative COVID-19 Vaccine Doses, by Age and Sex, in Adults Aged 65 Years and Older in the Medicare Shared Systems Data, by Vaccine Brand, Dec 11, 2020 to January 15, 2022.

Comparing data cuts through April 24, 2021 (Table S6), March 13, 2021 (Table S7) and February 27, 2021 (Table S8), the populations differed for those who were vaccinated from mid December 2020 to February 2021 versus the later months. Pfizer vaccinees experienced more hospitalizations in the prior year and higher proportions of underlying medical conditions (e.g., Charlson comorbidity index > 0), when compared to the overall elderly Medicare, Moderna, or Janssen populations.

### PMaxSPRT Sequential Testing Results

All outcomes with primary analyses for PMaxSPRT testing met the prespecified criteria for initiation of analyses (Table S3). No statistical signals were identified following vaccination with either the Moderna or Janssen vaccines. AMI (RR=1.42), PE (RR=1.54), disseminated intravascular coagulation (DIC; RR=1.91), and immune thrombocytopenia (ITP; RR=1.44) following Pfizer-BioNTech vaccination met the statistical threshold for a signal (Table 2). Dose-specific results can be seen in Tables S4-5.

**Table 2:** Summary of Sequential Testing Results in Adults Aged 65 years and Older in the Medicare Shared Systems Database for Any Dose by Vaccine Brand, Dec 11, 2020 to January 15, 2022.

| Outcomes, by Vaccine Brand | Vaccine Brand | Observed Person Time (Days) | Number of Doses | Number of Observed Outcomes (as of 01/15/2022)<sup>a</sup> | Relative Risk of Observed vs. Expected (Any Dose; 01/15/2022)<sup>b</sup> | Signal Identified <sup>c</sup> |
| --- | --- | --- | --- | --- | --- | --- |
| Acute Myocardial Infarction | Pfizer-BioNTech | 402,380,589 | 15,747,074 | 13,293 | 0.97 | Yes - 2/27/2021 (RR=1.42) |
|  | Moderna | 423,244,936 | 15,628,229 | 12,909 | 0.94 | No |
|  | Janssen | 13,156,815 | 570,434 | 548 | 1.30 | No |
| Deep Vein Thrombosis | Pfizer-BioNTech | 402,057,021 | 15,600,926 | 12,871 | 0.88 | No |
|  | Moderna | 424,775,904 | 15,507,286 | 11,342 | 0.78 | No |
|  | Janssen | 13,126,819 | 567,190 | 480 | 1.07 | No |
| Pulmonary Embolism | Pfizer-BioNTech | 404,173,653 | 15,684,098 | 9,443 | 1.15 | Yes – 2/27/2021 (RR=1.54) |
|  | Moderna | 426,520,976 | 15,571,477 | 8,996 | 1.08 | No |
|  | Janssen | 13,184,317 | 569,676 | 346 | 1.34 | No |
| Disseminated Intravascular Coagulation | Pfizer-BioNTech | 405,993,894 | 15,891,008 | 355 | 0.92 | Yes – 3/13/2021 (RR=1.91) |
|  | Moderna | 426,696,226 | 15,757,044 | 303 | 0.80 | No |
|  | Janssen | 13,297,825 | 576,495 | 14 | 1.11 | No |
| Non-hemorrhagic Stroke | Pfizer-BioNTech | 403,683,932 | 15,799,026 | 7,882 | 0.85 | No |
|  | Moderna | 424,648,679 | 15,680,668 | 7,646 | 0.83 | No |
|  | Janssen | 13,214,072 | 572,972 | 297 | 1.06 | No |
| Hemorrhagic Stroke | Pfizer-BioNTech | 405,580,548 | 15,874,528 | 2,128 | 0.96 | No |
|  | Moderna | 426,350,816 | 15,744,161 | 1,958 | 0.88 | No |
|  | Janssen | 13,283,534 | 575,871 | 77 | 1.12 | No |
| Immune Thrombocytopenia | Pfizer-BioNTech | 554,443,925 | 15,857,949 | 1,670 | 1.26 | Yes – 4/24/2021 (RR=1.44) |
|  | Moderna | 573,697,153 | 15,725,918 | 1,526 | 1.14 | No |
|  | Janssen | 19,848,219 | 575,565 | 62 | 1.34 | No |
| Myocarditis/Pericarditis | Pfizer-BioNTech | 555,100,577 | 15,876,623 | 1,415 | 1.10 | No |
|  | Moderna | 574,346,394 | 15,743,573 | 1,259 | 0.97 | No |
|  | Janssen | 19,865,485 | 576,075 | 48 | 1.04 | No |
| Guillain-Barre Syndrome | Pfizer-BioNTech | 532,555,069 | 15,892,318 | 69 | 1.14 | No |
|  | Moderna | 536,703,703 | 15,758,009 | 53 | 0.86 | No |
|  | Janssen | 19,582,979 | 576,548 | ** | 3.85 | No |
| Bell's Palsy | Pfizer-BioNTech | 575,124,693 | 16,453,363 | 3,618 | 1.12 | No |
|  | Moderna | 594,439,852 | 16,299,826 | 3,505 | 1.07 | No |
|  | Janssen | 20,810,662 | 603,113 | 133 | 1.13 | No |
| Encephalomyelitis/Encephalitis | Pfizer-BioNTech | 561,415,207 | 16,479,530 | 153 | 1.21 | No |
|  | Moderna | 572,296,487 | 16,324,225 | 120 | 0.98 | No |
|  | Janssen | 20,648,928 | 604,024 | ** | 1.68 | No |
| Transverse Myelitis | Pfizer-BioNTech | 541,656,398 | 15,892,229 | 66 | 1.36 | No |
|  | Moderna | 552,699,497 | 15,757,865 | 48 | 1.00 | No |
|  | Janssen | 19,700,796 | 576,539 | ** | 1.10 | No |
| Narcolepsy | Pfizer-BioNTech | 555,189,780 | 15,879,173 | 623 | 1.15 | No |
|  | Moderna | 574,433,145 | 15,745,902 | 537 | 0.98 | No |
|  | Janssen | 19,864,965 | 576,089 | 23 | 1.15 | No |
| Appendicitis | Pfizer-BioNTech | 548,982,786 | 15,880,177 | 1,180 | 1.06 | No |
|  | Moderna | 564,851,514 | 15,745,889 | 1,190 | 1.04 | No |
|  | Janssen | 19,785,242 | 576,111 | 37 | 0.86 | No |

<sup>a</sup> \*\* indicates small cell size counts with fewer than 11.
<sup>b</sup> The relative risk incorporates adjustments for observation delay as well as standardization by nursing home residency, sex, age, and race when indicated by in Table 3.

### Signal Evaluation

None of the prespecified data quality assurance checks, including claims duplication and unusual variability in claim accrual, raised data quality concerns (Table S9). Primary findings for signal robustness and signal characterization analyses are summarized in Table 3. Adjustment for monthly variation in the background rates resulted in statistically non-significant associations for AMI, DIC, and ITP following Pfizer-BioNTech vaccination. With background rates from the flu-vaccinated population as the historical comparator, DIC and ITP no longer met the signal threshold, while signals for AMI (RR=1.41) and PE (RR=1.48) remained. When rates during the peri-COVID period were used as the historical comparator, PE and DIC no longer met the signal threshold. We conducted an additional ad hoc sensitivity analysis for PE. When PE events were restricted to the inpatient setting, the statistical signal remained (RR=2.17).

**Table 3:**
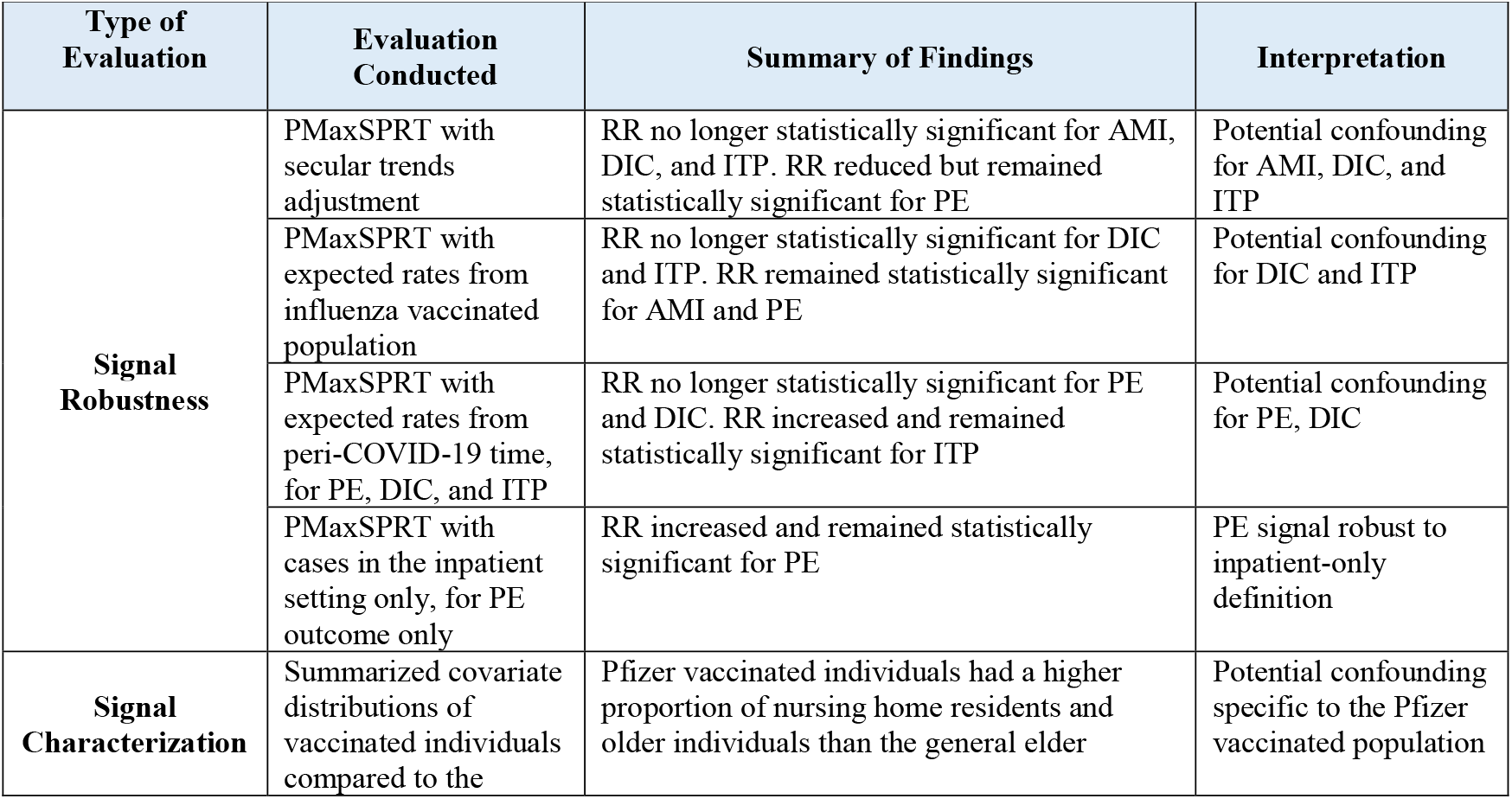
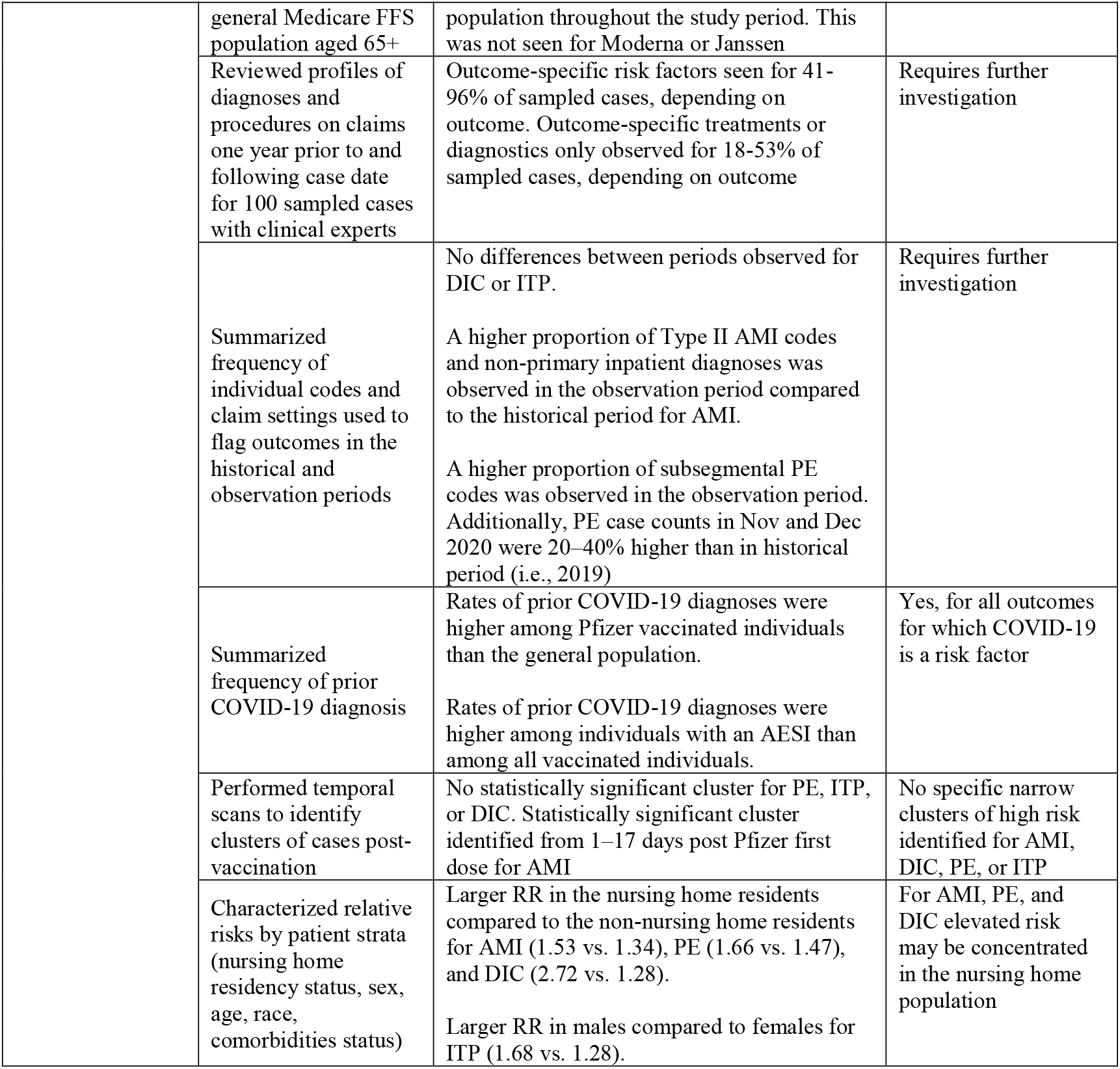
Summary of Characterization of Associations for Acute Myocardial Infarction, Pulmonary Embolism, Disseminated Intravascular Coagulation, and Immune Thrombocytopenia identified via Diagnosis Codes after Pfizer-BioNTech vaccination in Medicare Shared Systems Database.

Clinical subject matter experts reviewed claims-based diagnoses and procedures for selected patients from one year prior through one year after the dates of sampled outcome events. They found outcome-specific comorbidities were present in 41%, 45%, 95%, and 66% of AMI, PE, DIC, and ITP cases, respectively. Additionally, only a single diagnosis without other mention of the outcome occurred in 26%, 37%, 61%, and 49% of cases for AMI, PE, DIC, and ITP, respectively. Outcome-specific treatments or diagnostics were observed in 34%, 53%, 40%, and 18% of AMI, PE, DIC, and ITP cases, respectively.

Evaluation of claims-based diagnosis codes showed differences in coding patterns for AMI and PE between the study period and the historical period used to calculate expected rates (Table 3). Type II AMI codes, which are indicative of a mismatch between myocardial oxygen supply and demand (as opposed to acute coronary thrombosis), were more common in outcomes identified during the study period (46%) than in the historical period (28%). Additionally, for both AMI and PE outcomes, a higher proportion of inpatient claims-based codes occurred in a non-primary diagnosis position in the study period. No differences in coding patterns between the study period and historical period were noted for either ITP or DIC.

Finally, temporal scans conducted at the time of signal did not identify any clustering of cases within the post-vaccination risk window for PE, DIC, or ITP post Pfizer’s first dose. A scan for clusters 1-18 days long identified a statistically significant cluster of 1-17 days for AMI; however, the observed rate of events in the cluster was only 4% higher than if events had been evenly distributed in the risk window (Figure 2). Another scan for clusters 1-10 days long that also accounted for censoring due to end of follow-up did not identify a statistically significant cluster for AMI (Figure 2).

**Fig 2.**
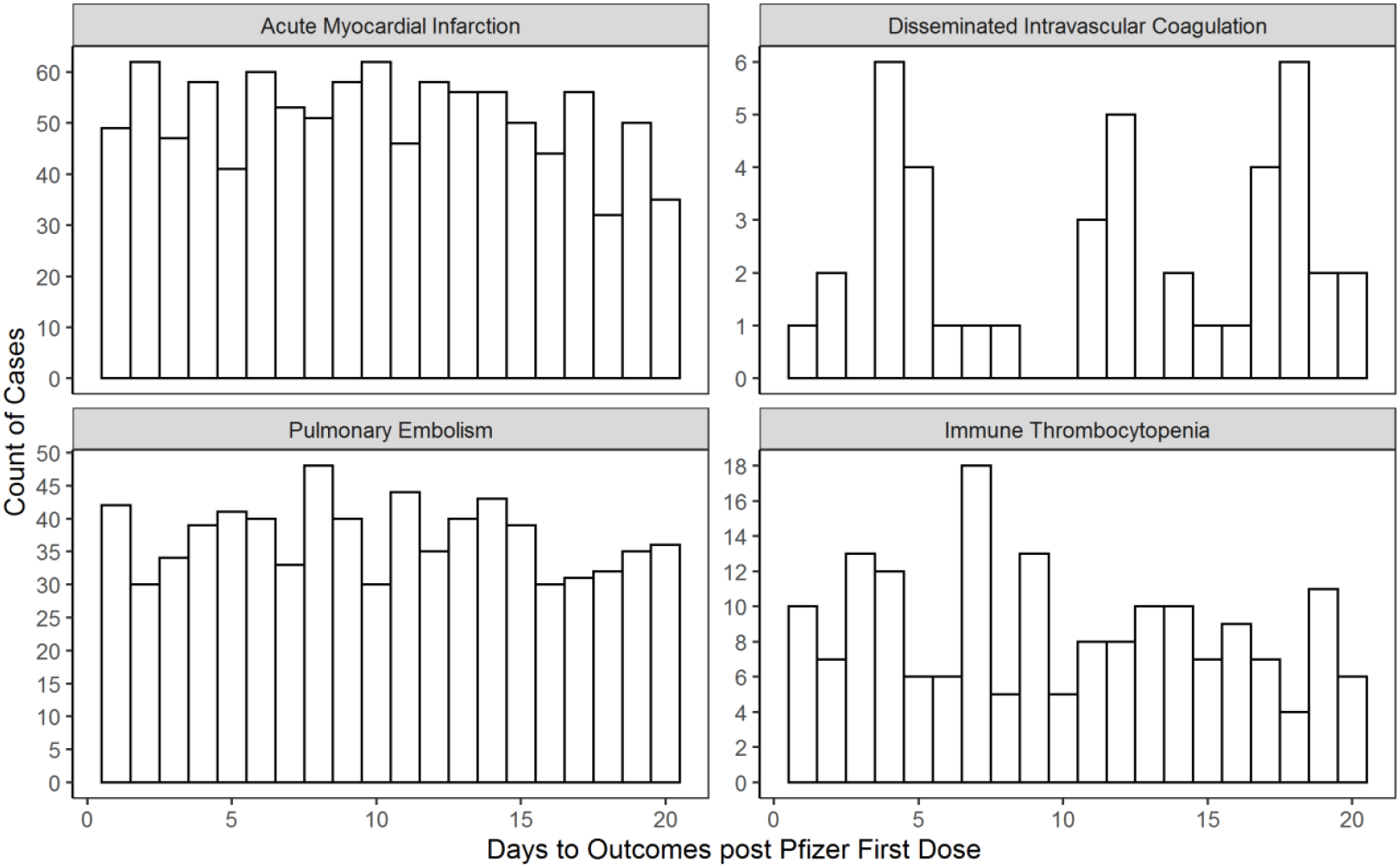
Distribution of Days to Diagnosis of Acute Myocardial Infarction, Pulmonary Embolism, Disseminated Intravascular Coagulation, or Immune Thrombocytopenia in 20 days after Pfizer-BioNTech Vaccination (First Dose) in Adults Aged 65 Years and Older, the Medicare Shared Systems Database.

## Discussion

Our early warning safety system is the first to identify four new statistical signals for modestly elevated risks (RR < 2) of four serious outcomes of AMI, PE, DIC, and ITP following Pfizer-BioNTech vaccination. This FDA and CMS COVID-19 vaccine safety study is one of the largest studies of elderly persons aged 65 years and above including approximately 34 million doses administered to more than 17 million Medicare insured persons. Our surveillance monitoring did not detect statistical signals for the Moderna and Janssen vaccines for any of the 14 monitored outcomes.

The statistical signals of four serious outcomes are not necessarily causal and may be due to factors potentially unrelated to vaccination. Additional analyses indicated that the potential association was less than twice the historical rates and may be associated with factors not accounted for in the near real-time surveillance methods. For example, the elderly Medicare population that received the Pfizer-BioNTech vaccine differed from other elderly COVID-19 vaccinated populations, including a preponderance of nursing home residents and populations with a higher comorbidity burden. These demographic and medical differences were not fully accounted for, since expected rates were only standardized to a subset of characteristics – age, sex, race, and nursing home residency status. Further, the AMI, DIC, and ITP signals were not robust when additional baseline rates were evaluated, while the PE signal might be explained by differences in rates between the pre-COVID-19 and peri-COVID-19 periods. In addition, the clinical assessment of patterns of reimbursement codes indicated that a substantial fraction had pre-existing outcome-specific comorbidities and risk factors, and that some outcomes may be due to follow-up care to an existing condition preceding the vaccination.

Our study has several strengths. This is the largest study of a population of more than 25 million elderly persons who are vulnerable to COVID-19 infections and complications-including residents of long-term care facilities. By using the large Medicare nationwide database with longitudinal linkage of vaccination, health services, and demographic information for millions of elderly persons, we can detect even small increases in the relative risk of rare outcomes for multiple vaccines that may not be captured in pre-authorization clinical trials. In addition, this near real-time surveillance benefits from the experience and knowledge obtained during more than a dozen years of successful collaboration between FDA and CMS conducting vaccine safety analyses using the Medicare database [13], including near real-time surveillance analyses for Guillain-Barré syndrome after influenza vaccination [14-16]. Furthermore, the weekly data updates and analyses allow for signal detection across 14 outcomes using near-real time monitoring. This further expands our knowledge of COVID-19 vaccine safety for informing timely regulatory action, if warranted as well as decision-making by healthcare providers, patients and the general public

We acknowledge our analysis has limitations. The near real-time analysis did not adjust for underlying risk factors such as comorbidities among recipients in the early vaccination campaign leading to falsely positive or negative signals. Furthermore, the early warning system may falsely identify a signal (false positive) or signals because of the high number of statistical tests performed or possible misspecification of parameters. Conversely, true safety signals (false negatives) may be missed due to mispecified parameters in the analyses. Diagnosis billing codes in claims data may underestimate or overestimate certain clinical conditions because of reimbursement priorities. We also note that results of this near real-time surveillance in elderly persons may not be generalizable to those younger than 65 years and adults who are uninsured or received only commercial health insurance. To address several of these limitations we are conducting further epidemiological studies along with medical record review to adjudicate outcomes identified by claims-based definitions.

In conclusion, we demonstrate that this FDA-CMS early warning safety system is working to rapidly identify potential new and important safety concerns following COVID-19 vaccination for consideration and to support potential decision-making by regulatory and public health authorities, healthcare professionals and the general public. Our new findings of statistical signals for four important outcomes for the Pfizer-BioNTech vaccine should be interpreted cautiously because the early warning system does not prove that vaccines cause the safety outcomes. FDA strongly believes the potential benefits of COVID-19 vaccination outweigh the potential risks of COVID-19 infection. Per FDA communication of these findings, FDA is currently not taking any regulatory actions based on these signal detection activities because these signals are still under investigation and require more robust study. The FDA active surveillance systems, including the CMS partnership, are a major part of a larger US federal surveillance effort to increase knowledge of COVID-19 vaccine safety to support decision-making that further protects public health during the pandemic.

## Data Availability

The study protocol was publicly posted as referenced in the manuscript before data analyses and related documents can be made available where needed, by contacting the corresponding author. De-identified participant data will not be shared without approval from CMS.

## Acknowledgements

We thank Joyce Obidi, PhD, Kristin “Kristine” Sepúlveda, MBA and Tainya Clarke, PhD, MPH, MSc for program management and J. Rosser Matthews, PhD, MPP, MPH for writing support. We would like to acknowledge John Hornberger, MD, MS, Nirmal Choradia, MD, Marna Bogan, RN, CPC, Susan Siford, PharmD, MBA, Christina Jessee, RN, CPC for their clinical and coding expertise. Additionally, we acknowledge Anchi Lo, MS, MPH, Jing Wang, BA, Yue Wu, MS, Zhiruo Wan, MSE, Shanlai Shangguan, MPH, Rowan McEvoy, BS, Arnstein Lindaas, MA, and Chianti Shi, MS for providing statistical programming support, and Yixin Jiao, MPP, Manzi Ngaiza, MPH, and Ellie Smith, BS for providing writing support. We acknowledge Michael Sklar, PhD, Lorene Nelson, PhD, MS, Julia Simard, ScD, SM, and Steve Goodman, MD, MHS, PhD for their epidemiologic expertise. Finally, we acknowledge Ivair Silva, PhD for his methodological expertise and development of the ‘Sequential’ R package used to conduct analyses in this manuscript.

## Notes

**Conflict of Interest Statement** All authors have completed the ICMJE uniform disclosure form at www.icmje.org/coi_disclosure.pdf and declare: Hui-Lee Wong, Cindy Zhou, Deborah Thompson, Rositsa Dimova, Patricia Lloyd, Richard Forshee, and Steven Anderson are/were salaried employees of the U.S. Food and Drug Administration; Ellen Tworkoski, Mao Hu, Bradley Lufkin, Rose Do, Laurie Feinberg, Yoganand Chillarige, and Thomas MaCurdy are/were salaried employees of Acumen, LLC, which is a contractor with the U.S. Food and Drug Administration; Jeffrey Kelman is a salaried employee of the Centers for Medicare & Medicaid Services; Ellen Tworkoski is currently a salaried employee of Biogen, which includes stock options provided to her as part of employee benefits; no other relationships or activities that could appear to have influenced the submitted work.

### Competing Interest Statement

All authors have completed the ICMJE uniform disclosure form at www.icmje.org/coi_disclosure.pdf and declare: Hui-Lee Wong, Cindy Zhou, Deborah Thompson, Rositsa Dimova, Patricia Lloyd, Richard Forshee, and Steven Anderson are/were salaried employees of the U.S. Food and Drug Administration; Ellen Tworkoski, Mao Hu, Bradley Lufkin, Rose Do, Laurie Feinberg, Yoganand Chillarige, and Thomas MaCurdy are/were salaried employees of Acumen, LLC, which is a contractor with the U.S. Food and Drug Administration; Jeffrey Kelman is a salaried employee of the Centers for Medicare & Medicaid Services; Ellen Tworkoski is currently a salaried employee of Biogen, which includes stock options provided to her as part of employee benefits; no other relationships or activities that could appear to have influenced the submitted work.

### Funding Statement

The US Food and Drug Administration provided funding for this study and contributed as follows: led the design of the study, interpretation of the results, writing of the manuscript, decision to submit, and made contributions to the coordination of data collection and analysis of the data.

### Author Declarations

We conducted this study within the Biologics Effectiveness and Safety (BEST) Initiative. FDA Amendments Act of 2007 required that FDA develop a national electronic system for monitoring safety of FDA-regulated medical products. FDA built the Sentinel Initiative in response to the congressional requirement, and the BEST Initiative commenced as a Center for Biologics Evaluation and Research (CBER) component of Sentinel. The Office of Human Research Protection (OHRP) determined that the regulations OHRP administers (45 CFR part 46) do not apply to the activities that are included in the FDA's Sentinel Initiative.

