## Supplemental File 1 for "Surveillance of COVID-19 vaccine safety among elderly persons aged 65 years and older"

**Supplemental Tables and Figures**

Table S1. Codes for COVID-19 Vaccines

Table S2. Characteristics of Prespecified Potential Adverse Events of Interests (AESI)* in Sequential Hypothesis Testing

Table S3. Prespecified Parameters for PMaxSPRT Sequential Hypothesis Testing

Table S4: Summary of Sequential Testing Results in Adults Aged 65 years and Older in the Medicare Shared Systems Database for Dose 2 by Vaccine Brand, Dec 11, 2020 to January 15, 2022

Table S5: Summary of Sequential Testing Results in Adults Aged 65 years and Older in the Medicare Shared Systems Database for Dose 2 by Vaccine Brand, Dec 11, 2020 to January 15, 2022

Table S6. Characteristics of Pfizer-BioNTech, Moderna, Janssen Vaccine Doses Administered among Adults Aged 65 years and Older in Medicare[a], Dec 11, 2020 to Apr 24, 2021

Table S7. Characteristics of Pfizer-BioNTech, Moderna, Janssen Vaccine Doses Administered among Adults Aged 65 years and Older in Medicare[a], Dec 11, 2020 to Mar 13, 2021

Table S8. Characteristics of Pfizer-BioNTech and Moderna Vaccine Doses[a] Administered among Adults Aged 65 years and Older in Medicare[b], Dec 11, 2020 to Feb 27, 2021

Table S9. Summary of Data Quality Checks for Acute Myocardial Infarction, Pulmonary Embolism, Disseminated Intravascular Coagulation, and Immune Thrombocytopenia identified via Diagnosis Codes after Pfizer-BioNTech vaccination in Medicare Shared Systems Database

Table S10. Summary of Primary and Sensitivity Rapid Cycle Analyses for Acute Myocardial Infarction, Pulmonary Embolism, Disseminated Intravascular Coagulation, and Immune Thrombocytopenia identified after Pfizer-BioNTech vaccination in Medicare Shared Systems Database

**Table S1. Codes for COVID-19 Vaccines**

| Manufacturer | Name | HCPCS/CPT Code[1] | Vaccine Administration Code | Dosing Interval |
|---|---|---|---|---|
| Pfizer | Pfizer-BioNTech COVID-19 Vaccine | 91300 | 0001A (1st dose) | - 21 Days between doses 1, 2<br>- 28 Days between doses 2, 3<br>- 6 months between primary series and booster dose |
| | | | 0002A (2nd dose) | |
| | | | 0003A (3rd dose) | |
| | | | 0004A (Booster dose) | |
| | Pfizer-BioNTech COVID-19 Vaccine (Ready to Use) | 91305 | 0051A (1st dose, ready to use) | |
| | | | 0052A (2nd dose, ready to use) | |
| | | | 0053A (3rd dose, ready to use) | |
| | | | 0054A (Booster dose, ready to use) | |
| Moderna | Moderna COVID-19 Vaccine | 91301 | 0011A (1st dose) | - 28 Days between doses 1, 2<br>- 28 Days between doses 2, 3<br>- 6 months between primary series and booster dose |
| | | | 0012A (2nd dose) | |
| | | | 0013A (3rd dose) | |
| | Moderna COVID-19 Vaccine (Low Dose) | 91306 | 0064A (low-dose booster) | |
| Janssen | Janssen COVID-19 Vaccine | 91303 | 0031A | - 2 months between primary and vaccination and booster dose |
| | | | 0034A (booster) | |

Abbreviations: CPT, Current Procedural Terminology, HCPCS, Healthcare Common Procedure Coding System

### Table S2. Characteristics of Prespecified Potential Adverse Events of Interests (AESI)* in Sequential Hypothesis Testing

| AESI | Rationale for Inclusion as Prespecified AESI[a] | Setting[b] | Clean Window[c] | Risk Window |
|---|---|---|---|---|
| **Vascular/Hematologic** | | | | |
| Acute Myocardial Infarction | 1 | IP | 365 days | 1-28 days[2,3] |
| Deep Vein Thrombosis | 1 | IP, OP/PB | 365 days | 1-28 days[4-6] |
| Pulmonary Embolism[d] | 1 | IP, OP/PB | 365 days | 1-28 days[4,5,7] |
| Disseminated Intravascular Coagulation | 1 | IP, OP-ED | 365 days | 1-28 days[8] |
| Non-hemorrhagic Stroke | 1 | IP | 365 days | 1-28 days[2,3] |
| Hemorrhagic Stroke | 1 | IP | 365 days | 1-28 days[2,3] |
| Immune Thrombocytopenia | 2 | IP, OP/PB | 365 days | 1-42 days[9,10] |
| **Cardiac (Non-Vascular)** | | | | |
| Myocarditis/Pericarditis | 2 | IP, OP/PB | 365 days | 1-42 days[11] |
| **Neurological** | | | | |
| Guillain-Barré Syndrome | 2 | IP- primary position only | 365 days | 1-42 days[12,13] |
| Bell's Palsy | 2 | IP, OP/PB | 183 days | 1-42 days[14] |
| Encephalomyelitis/Encephalitis | 2 | IP | 183 days | 1-42 days[15] |
| Transverse Myelitis | 2 | IP, OP-ED | 365 days | 1-42 days[16] |
| Narcolepsy | 4 | IP, OP/PB | 365 days | 1-42 days[e],[17-19] |
| **Gastrointestinal** | | | | |
| Appendicitis | 4 | IP, OP-ED | 365 days | 1-42 days[7,20] |

*These AESI have not been associated with COVID-19 vaccines based on available pre-licensure evidence.

[a] Categories for Inclusion in the Prespecified adverse events of special interest (AESI) list

1. Possible enhanced disease following immunization
2. Theoretical concerns based on associations with other vaccines (e.g., influenza vaccines)
3. Passive surveillance from similar vaccine platforms
4. Others including recommendations from other vaccine surveillance systems

[b] Setting Definitions: IP refers to inpatient facility claims. OP-ED refers to a subset of outpatient facility claims occurring in the emergency department. OP/PB refers to all outpatient facility claims, and professional/provider claims except those professional/provider claims with a laboratory place of service.

[c] References for the duration of these windows could not be located in the literature and are instead based on input from clinicians.

[d] If an individual has both deep vein thrombosis (DVT) and pulmonary embolism (PE) (i.e., the DVT progressed to PE), the case will be de-duplicated in analyses stage and assigned only PE. The PE onset date is determined by the date the PE code is reported in the database.

[e] Literature typically uses a longer window duration, but we propose a shorter risk period for the purposes of rapid signal detection, assuming that risk should either be constant or more concentrated in a shorter window nearer to the time of vaccination.

Note: MIS and anaphylaxis are not in the formal sequential testing analyses as of May 1, 2021. Only descriptive statistics were generated.

Definitions: **Clean Window** is defined as an interval used to define incident outcomes where an individual enters the study cohort only if the AESI of interest did not occur during that interval. **Risk Window** is defined as an interval during which occurrence of the AESI of interest will be included in the analyses. **Adverse event of special interest (AESI)** is defined as "[A]n adverse event of special interest (serious or non-serious) is one of scientific and medical concern specific to the sponsor's product or program, for which ongoing monitoring and rapid communication by the investigator to the sponsor could be appropriate. Such an event might require further investigation in order to characterize and understand it. Depending on the nature of the event, rapid communication by the trial sponsor to other parties (e.g., regulators) might also be warranted." Source: The Development Safety Update Report: Harmonizing the Format and Content for Periodic Safety Reporting During Clinical Trials: Report of Council for International Organizations of Medical Sciences Working Group VII, Geneva 2007. https://cioms.ch/shop/product/development-safety-update-report-dsur-harmonizing-format-contentperiodic-safety-report-clinical- trials-report-cioms-working-group-vii/ (last accessed May 2021)

### Table S3. Pre-specified Parameters for PMaxSPRT Sequential Hypothesis Testing

| AESI | Study Population for Background Rates [21,22] | Annual Background Rates | | Testing Margin |
|---|---|---|---|---|
| | | Overall Rate per 100,000 person-years | RCA Stratifications | |
| Acute Myocardial Infarction | General Age 65+, 2018 | 1262.68 | NH*age*sex*race | 1.25 |
| Deep Vein Thrombosis | General Age 65+, 2019 | 1325.45 | NH*age*sex*race | 1.25 |
| Pulmonary Embolism | General Age 65+, 2019 | 751.68 | NH*age*sex*race | 1.25 |
| Disseminated Intravascular Coagulation | General Age 65+, 2019 | 36.90 | NH*age*sex*race | 1.25 |
| Non-hemorrhagic Stroke | General Age 65+, 2019 | 842.29 | NH*age*sex*race | 1.25 |
| Hemorrhagic Stroke | General Age 65+, 2019 | 205.18 | NH*age*sex*race | 1.25 |
| Immune Thrombocytopenia | General Age 65+, 2019 | 89.61 | NH*age*sex | 1.25 |
| Myocarditis/Pericarditis | General Age 65+, 2019 | 88.00 | NH*age*sex | 1.5 |
| Guillain-Barré Syndrome | General Age 65+, 2018 | 4.45 | NH | 2.5 |
| Bell's Palsy | General Age 65+, 2019 | 214.10 | NH*age*sex*race | 1.25 |
| Encephalomyelitis/ Encephalitis | General Age 65+, 2017 | 8.65 | NH*age | 2.5 |
| Transverse Myelitis | General Age 65+, 2017 | 3.39 | NH*age | 1.5 |
| Narcolepsy | General Age 65+, 2017 | 37.24 | NH*age*sex | 2.5 |
| Appendicitis | General Age 65+, 2019 | 80.06 | NH*age*sex | 1.25 |

Other PMaxSPRT Parameters:

- Testing frequency: Weekly
- Minimum # of events to signal = 3
- Total alpha per analysis: 1%
- Alpha spending plan: Constant
- Claims delay: Estimated from events occurring from Jan-Dec 2019
- Critical Value: Calculated from the R package 'Sequential'
- Maximum surveillance length: Number of events expected during the vaccination campaign based on anticipated # of vaccine doses administered over one year;
- Testing margins are defined based on comparator rates (from historical data) of the outcome θ adjusted for the length of the risk window and a target number of doses needed to harm [23]

**Table S4: Summary of Sequential Testing Results in Adults Aged 65 years and Older in the Medicare Shared Systems Database for Dose 1 by Vaccine Brand, Dec 11, 2020 to January 15, 2022**

| Outcomes, by Vaccine Brand | Vaccine Brand | Observed Person Time (Days) | Number of Doses | Number of Observed Outcomes (as of 01/15/2022)[a] | Relative Risk of Observed vs. Expected (Any Dose; 01/15/2022)[b] | Signal Identified [c] |
|---|---|---|---|---|---|---|
| Acute Myocardial Infarction | Pfizer-BioNTech | 114,975,224 | 5,151,154 | 4,519 | 1.08 | Yes - 2/27/2021 (RR=1.43) |
| | Moderna | 143,113,654 | 5,182,588 | 4,767 | 0.97 | |
| | Janssen | 13,156,815 | 475,817 | 548 | 1.30 | |
| Deep Vein Thrombosis | Pfizer-BioNTech | 114,001,357 | 5,103,509 | 4,302 | 0.97 | |
| | Moderna | 142,122,801 | 5,143,344 | 4,142 | 0.80 | |
| | Janssen | 13,126,819 | 473,240 | 480 | 1.07 | |
| Pulmonary Embolism | Pfizer-BioNTech | 114,670,124 | 5,133,709 | 3,203 | 1.31 | Yes - 2/27/2021 (RR=1.59) |
| | Moderna | 142,735,189 | 5,165,643 | 3,374 | 1.15 | |
| | Janssen | 13,184,317 | 475,325 | 346 | 1.34 | |
| Disseminated Intravascular Coagulation | Pfizer-BioNTech | 116,150,215 | 5,203,952 | 130 | 1.07 | |
| | Moderna | 144,377,720 | 5,228,712 | 122 | 0.86 | |
| | Janssen | 13,297,825 | 480,992 | 14 | 1.11 | |
| Non-hemorrhagic Stroke | Pfizer-BioNTech | 115,382,015 | 5,169,392 | 2,488 | 0.88 | |
| | Moderna | 143,605,453 | 5,200,556 | 2,807 | 0.85 | |
| | Janssen | 13,214,072 | 477,929 | 297 | 1.06 | |
| Hemorrhagic Stroke | Pfizer-BioNTech | 116,010,479 | 5,197,652 | 686 | 1.01 | |
| | Moderna | 144,248,107 | 5,223,980 | 727 | 0.90 | |
| | Janssen | 13,283,534 | 480,468 | 77 | 1.12 | |
| Immune Thrombocytopenia | Pfizer-BioNTech | 123,092,403 | 5,193,253 | 418 | 1.36 | |
| | Moderna | 154,716,873 | 5,218,709 | 433 | 1.14 | |
| | Janssen | 19,848,219 | 480,249 | 62 | 1.34 | |
| Myocarditis/Pericarditis | Pfizer-BioNTech | 123,236,370 | 5,199,277 | 352 | 1.16 | |
| | Moderna | 154,882,448 | 5,224,251 | 368 | 0.99 | |
| | Janssen | 19,865,485 | 480,665 | 48 | 1.04 | |
| Guillain-Barre Syndrome | Pfizer-BioNTech | 122,943,541 | 5,204,450 | 19 | 1.30 | |
| | Moderna | 154,504,965 | 5,229,054 | 16 | 0.85 | |
| | Janssen | 19,582,979 | 481,041 | ** | 3.85 | |
| Bell's Palsy | Pfizer-BioNTech | 127,726,357 | 5,387,266 | 837 | 1.11 | |
| | Moderna | 160,409,091 | 5,410,935 | 1,024 | 1.09 | |
| | Janssen | 20,810,662 | 503,719 | 133 | 1.13 | |
| Encephalomyelitis/Encephalitis | Pfizer-BioNTech | 127,656,653 | 5,396,075 | 35 | 1.11 | |
| | Moderna | 160,313,295 | 5,419,104 | 37 | 0.97 | |
| | Janssen | 20,648,928 | 504,483 | ** | 1.68 | |
| Transverse Myelitis | Pfizer-BioNTech | 123,095,510 | 5,204,417 | 16 | 1.34 | |
| | Moderna | 154,702,192 | 5,229,022 | 16 | 1.11 | |
| | Janssen | 19,700,796 | 481,033 | ** | 1.10 | |

| Outcomes, by Vaccine Brand | Vaccine Brand | Observed Person Time (Days) | Number of Doses | Number of Observed Outcomes (as of 01/15/2022)[a] | Relative Risk of Observed vs. Expected (Any Dose; 01/15/2022)[b] | Signal Identified [c] |
|---|---|---|---|---|---|---|
| Narcolepsy | Pfizer-BioNTech | 123,255,702 | 5,200,093 | 155 | 1.21 | |
| | Moderna | 154,904,510 | 5,225,006 | 169 | 1.07 | |
| | Janssen | 19,864,965 | 480,652 | 23 | 1.15 | |
| Appendicitis | Pfizer-BioNTech | 123,146,290 | 5,200,623 | 284 | 1.09 | |
| | Moderna | 154,760,041 | 5,225,114 | 299 | 0.90 | |
| | Janssen | 19,785,242 | 480,672 | 37 | 0.86 | |

[a] ** indicates small cell size counts with fewer than 11.

[b] The relative risk incorporates adjustments for observation delay as well as standardization by nursing home residency, sex, age, and race when indicated by in Table 3.

**Table S5: Summary of Sequential Testing Results in Adults Aged 65 years and Older in the Medicare Shared Systems Database for Dose 2 by Vaccine Brand, Dec 11, 2020 to January 15, 2022**

| Outcomes, by Vaccine Brand | Vaccine Brand | Observed Person Time (Days) | Number of Doses | Number of Observed Outcomes (as of 01/15/2022)[a] | Relative Risk of Observed vs. Expected (Any Dose; 01/15/2022)[b] | Signal Identified[c] |
|---|---|---|---|---|---|---|
| Acute Myocardial Infarction | Pfizer-BioNTech | 129,180,459 | 4,633,937 | 4,546 | 0.97 | Yes - 3/6/2021 (RR=1.45) |
| | Moderna | 130,110,732 | 4,663,865 | 4,233 | 0.95 | |
| Deep Vein Thrombosis | Pfizer-BioNTech | 128,037,671 | 4,590,685 | 4,434 | 0.90 | |
| | Moderna | 129,162,959 | 4,628,141 | 3,734 | 0.80 | |
| Pulmonary Embolism | Pfizer-BioNTech | 128,755,392 | 4,616,589 | 3,143 | 1.15 | Yes - 3/13/2021 (RR=1.47) |
| | Moderna | 129,701,407 | 4,647,487 | 2,899 | 1.10 | |
| Disseminated Intravascular Coagulation | Pfizer-BioNTech | 130,420,051 | 4,678,849 | 133 | 0.99 | |
| | Moderna | 131,211,022 | 4,703,547 | 107 | 0.84 | |
| Non-hemorrhagic Stroke | Pfizer-BioNTech | 129,617,235 | 4,649,782 | 2,854 | 0.91 | |
| | Moderna | 130,545,761 | 4,679,558 | 2,633 | 0.88 | |
| Hemorrhagic Stroke | Pfizer-BioNTech | 130,273,915 | 4,673,550 | 755 | 0.99 | |
| | Moderna | 131,100,836 | 4,699,566 | 659 | 0.91 | |
| Immune Thrombocytopenia | Pfizer-BioNTech | 194,914,543 | 4,669,279 | 646 | 1.34 | |
| | Moderna | 196,189,048 | 4,694,544 | 602 | 1.26 | |
| Myocarditis/Pericarditis | Pfizer-BioNTech | 195,139,639 | 4,674,668 | 489 | 1.02 | |
| | Moderna | 196,399,564 | 4,699,580 | 445 | 0.95 | |
| Guillain-Barre Syndrome | Pfizer-BioNTech | 194,948,451 | 4,679,241 | 26 | 1.12 | |
| | Moderna | 196,288,336 | 4,703,848 | 24 | 1.01 | |
| Bell's Palsy | Pfizer-BioNTech | 202,125,663 | 4,842,563 | 1,337 | 1.12 | |
| | Moderna | 203,393,056 | 4,867,368 | 1,293 | 1.09 | |
| Encephalomyelitis/Encephalitis | Pfizer-BioNTech | 202,197,205 | 4,850,317 | 51 | 1.04 | |
| | Moderna | 203,503,561 | 4,874,546 | 44 | 0.92 | |
| Transverse Myelitis | Pfizer-BioNTech | 195,090,508 | 4,679,219 | 24 | 1.29 | |
| | Moderna | 196,396,121 | 4,703,808 | 21 | 1.16 | |
| Narcolepsy | Pfizer-BioNTech | 195,168,873 | 4,675,381 | 247 | 1.23 | |
| | Moderna | 196,426,852 | 4,700,238 | 205 | 1.04 | |
| Appendicitis | Pfizer-BioNTech | 195,076,445 | 4,675,833 | 455 | 1.10 | |
| | Moderna | 196,347,122 | 4,700,332 | 467 | 1.10 | |

[a] ** indicates small cell size counts with fewer than 11.

[b] The relative risk incorporates adjustments for observation delay as well as standardization by nursing home residency, sex, age, and race when indicated by in Table 3.

**Table S6. Characteristics of Pfizer-BioNTech, Moderna, Janssen Vaccine Doses Administered among Adults Aged 65 years and Older in Medicare[a], Dec 11, 2020 to Apr 24, 2021**

| Patient Characteristic | General 65+ FFS[b] | Moderna[c] | Pfizer | Janssen | SMD - Comparison with General FFS | | |
|---|---|---|---|---|---|---|---|
| | | | | | Moderna | Pfizer | Janssen |
| **Total** | **25,390,578** | **7,569,305** | **7,319,407** | **230,213** | | | |
| **Nursing Home Residential Status** | | | | | | | |
| Nursing Home Resident | 2.3% | 2.8% | 7.5% | 0.9% | 0.03 | **0.24** | **0.11** |
| Non-Nursing Home Resident | 97.7% | 97.2% | 92.5% | 99.1% | 0.03 | **0.24** | **0.11** |
| **Age (years)** | | | | | | | |
| 65-74 | 53.8% | 51.6% | 47.1% | 61.3% | 0.04 | **0.13** | **0.15** |
| 75-84 | 32.6% | 34.2% | 34.4% | 28.1% | 0.03 | 0.04 | 0.10 |
| 85+ | 13.5% | 14.2% | 18.5% | 10.6% | 0.02 | **0.14** | 0.09 |
| **Sex** | | | | | | | |
| Female | 55.9% | 57.8% | 59.8% | 57.8% | 0.04 | 0.08 | 0.04 |
| Male | 44.1% | 42.2% | 40.2% | 42.2% | 0.04 | 0.08 | 0.04 |
| **Race/Ethnicity** | | | | | | | |
| Asian | 2.1% | 2.0% | 1.9% | 2.1% | 0.01 | 0.01 | 0.00 |
| Black | 6.8% | 4.3% | 5.4% | 6.0% | **0.11** | 0.06 | 0.03 |
| Hispanic | 1.6% | 0.9% | 0.9% | 1.3% | 0.06 | 0.06 | 0.02 |
| Alaskan Native/Native American | 0.5% | 0.5% | 0.5% | 0.2% | 0.00 | 0.01 | 0.05 |
| White | 85.0% | 88.0% | 87.1% | 86.6% | 0.09 | 0.06 | 0.04 |
| Other | 1.8% | 1.8% | 1.8% | 1.4% | 0.00 | 0.00 | 0.03 |
| Missing/Unknown | 2.3% | 2.5% | 2.4% | 2.5% | 0.02 | 0.00 | 0.01 |
| **Urban/Rural** | | | | | | | |
| Urban | 76.5% | 75.5% | 84.0% | 75.9% | 0.02 | **0.19** | 0.01 |
| Rural | 22.9% | 24.0% | 15.6% | 23.8% | 0.03 | **0.19** | 0.02 |
| Missing/Unknown | 0.6% | 0.5% | 0.4% | 0.3% | 0.02 | 0.03 | 0.05 |
| **Medical Conditions (0-365 Days Prior to Index Date)** | | | | | | | |
| Atrial Fibrillation | 13.7% | 14.7% | 16.3% | 12.7% | 0.03 | 0.07 | 0.03 |
| AMI Hospitalization | 0.7% | 0.6% | 0.7% | 0.7% | 0.01 | 0.00 | 0.00 |
| Bronchiectasis | 0.9% | 1.0% | 1.1% | 0.8% | 0.01 | 0.02 | 0.01 |
| Charlson Comorbidity Index >0 | 63.3% | 65.8% | 69.1% | 63.3% | 0.05 | **0.12** | 0.00 |
| Asthma w/o COPD | 4.8% | 5.2% | 5.2% | 5.0% | 0.02 | 0.02 | 0.01 |
| COPD | 12.0% | 11.6% | 12.3% | 12.4% | 0.01 | 0.01 | 0.01 |
| Coronary Revascularization | 0.9% | 0.9% | 0.9% | 0.9% | 0.00 | 0.01 | 0.00 |
| Diabetes | 27.0% | 27.1% | 27.8% | 28.2% | 0.00 | 0.02 | 0.03 |
| Depression | 17.0% | 17.6% | 21.9% | 16.9% | 0.02 | **0.12** | 0.00 |

| Patient Characteristic | General 65+ FFS[b] | Moderna[c] | Pfizer | Janssen | SMD - Comparison with General FFS | | |
|---|---|---|---|---|---|---|---|
| | | | | | Moderna | Pfizer | Janssen |
| Gout | 5.4% | 5.7% | 5.7% | 5.3% | 0.01 | 0.01 | 0.00 |
| Hospitalization | 13.1% | 12.2% | 15.3% | 12.4% | 0.03 | 0.07 | 0.02 |
| Hypertension | 67.8% | 71.2% | 72.7% | 69.3% | 0.07 | **0.11** | 0.03 |
| Interstitial Lung Disease | 1.9% | 1.9% | 2.1% | 1.7% | 0.01 | 0.02 | 0.01 |
| Impaired Mobility | 0.4% | 0.4% | 0.7% | 0.3% | 0.01 | 0.04 | 0.01 |
| Neurological/Neurodevelopmental Conditions | 7.2% | 7.2% | 9.7% | 6.5% | 0.00 | 0.09 | 0.03 |
| Obesity | 20.9% | 21.7% | 20.6% | 22.3% | 0.02 | 0.01 | 0.03 |
| Pneumonia | 5.0% | 4.4% | 6.1% | 4.5% | 0.03 | 0.05 | 0.03 |
| Hospitalized Stroke | 0.6% | 0.6% | 0.8% | 0.6% | 0.01 | 0.01 | 0.01 |
| **Prior COVID-19 Diagnosis** | | | | | | | |
| Any | 3.3% | 4.6% | 7.0% | 6.0% | 0.07 | **0.17** | **0.13** |
| Inpatient | 0.9% | 1.0% | 1.6% | 1.5% | 0.01 | 0.06 | 0.05 |
| Outpatient or Professional | 3.3% | 4.6% | 7.0% | 5.9% | 0.07 | **0.17** | **0.13** |

Abbreviations: FFS, fee-for-service; SMD, standardized mean difference; AMI, acute myocardial infarction; COPD, chronic obstructive pulmonary disease; COVID-19, coronavirus disease 2019

[a] Individuals included in this table were required to have 365 days of continuous enrollment prior to vaccination date in order to accurately capture medical history

[b] Characteristics of the general Medicare FFS population aged 65 years and older were assessed as of 12/10/2020

[c] Characteristics of the vaccination doses (e.g., Moderna vaccine doses) were assessed at the administration date of the first dose of each vaccinated individual

**Table S7. Characteristics of Pfizer-BioNTech, Moderna, Janssen Vaccine Doses Administered among Adults Aged 65 years and Older in Medicare[a], Dec 11, 2020 to Mar 13, 2021**

| Patient Characteristic | General 65+ FFS[b] | Moderna[c] | Pfizer | Janssen | SMD - Comparison with General FFS | | |
|---|---|---|---|---|---|---|---|
| | | | | | Moderna | Pfizer | Janssen |
| **Total** | **25,390,578** | **5,846,688** | **5,636,731** | **104,753** | | | |
| **Nursing Home Residential Status** | | | | | | | |
| Nursing Home Resident | 2.3% | 3.4% | 9.6% | 0.3% | 0.07 | **0.31** | **0.18** |
| Non-Nursing Home Resident | 97.7% | 96.6% | 90.4% | 99.7% | 0.07 | **0.31** | **0.18** |
| **Age (years)** | | | | | | | |
| 65-74 | 53.8% | 49.1% | 42.4% | 64.6% | 0.10 | **0.23** | **0.22** |
| 75-84 | 32.6% | 35.7% | 36.5% | 26.1% | 0.06 | 0.08 | **0.14** |
| 85+ | 13.5% | 15.3% | 21.2% | 9.3% | 0.05 | **0.20** | **0.14** |
| **Sex** | | | | | | | |
| Female | 55.9% | 57.7% | 60.1% | 57.5% | 0.04 | 0.09 | 0.03 |
| Male | 44.1% | 42.3% | 39.9% | 42.5% | 0.04 | 0.09 | 0.03 |
| **Race/Ethnicity** | | | | | | | |
| Asian | 2.1% | 1.9% | 1.8% | 1.9% | 0.01 | 0.02 | 0.01 |
| Black | 6.8% | 4.1% | 4.9% | 5.4% | **0.12** | 0.08 | 0.06 |
| Hispanic | 1.6% | 0.8% | 0.8% | 1.3% | 0.08 | 0.07 | 0.02 |
| Alaskan Native/Native American | 0.5% | 0.5% | 0.6% | 0.1% | 0.01 | 0.02 | 0.06 |
| White | 85.0% | 88.4% | 87.9% | 87.1% | 0.10 | 0.09 | 0.06 |
| Other | 1.8% | 1.8% | 1.8% | 1.4% | 0.01 | 0.00 | 0.03 |
| Missing/Unknown | 2.3% | 2.5% | 2.2% | 2.8% | 0.01 | 0.01 | 0.03 |
| **Urban/Rural** | | | | | | | |
| Urban | 76.5% | 75.3% | 83.3% | 78.6% | 0.03 | **0.17** | 0.05 |
| Rural | 22.9% | 24.2% | 16.3% | 21.1% | 0.03 | **0.17** | 0.04 |
| Missing/Unknown | 0.6% | 0.5% | 0.4% | 0.3% | 0.02 | 0.03 | 0.05 |
| **Medical Conditions (0-365 Days Prior to Index Date)** | | | | | | | |
| Atrial Fibrillation | 13.7% | 15.2% | 17.5% | 12.2% | 0.04 | **0.10** | 0.04 |
| AMI Hospitalization | 0.7% | 0.6% | 0.8% | 0.6% | 0.01 | 0.01 | 0.01 |
| Bronchiectasis | 0.9% | 1.1% | 1.2% | 0.8% | 0.01 | 0.02 | 0.01 |
| Charlson Comorbidity Index >0 | 63.3% | 66.4% | 70.7% | 62.8% | 0.06 | **0.16** | 0.01 |
| Asthma w/o COPD | 4.8% | 5.2% | 5.1% | 5.3% | 0.02 | 0.02 | 0.02 |
| COPD | 12.0% | 11.5% | 12.7% | 11.5% | 0.01 | 0.02 | 0.02 |
| Coronary Revascularization | 0.9% | 0.9% | 0.9% | 0.9% | 0.00 | 0.01 | 0.00 |
| Diabetes | 27.0% | 26.9% | 27.8% | 27.6% | 0.00 | 0.02 | 0.01 |
| Depression | 17.0% | 17.8% | 23.2% | 17.2% | 0.02 | **0.16** | 0.01 |

| Patient Characteristic | General 65+ FFS[b] | Moderna[c] | Pfizer | Janssen | SMD - Comparison with General FFS | | |
|---|---|---|---|---|---|---|---|
| | | | | | Moderna | Pfizer | Janssen |
| Gout | 5.4% | 5.8% | 5.8% | 5.3% | 0.02 | 0.02 | 0.01 |
| Hospitalization | 13.1% | 12.4% | 16.3% | 11.2% | 0.02 | 0.09 | 0.06 |
| Hypertension | 67.8% | 71.7% | 73.7% | 69.0% | 0.08 | **0.13** | 0.03 |
| Interstitial Lung Disease | 1.9% | 2.0% | 2.2% | 1.7% | 0.01 | 0.02 | 0.01 |
| Impaired Mobility | 0.4% | 0.4% | 0.8% | 0.3% | 0.00 | 0.05 | 0.03 |
| Neurological/Neurodevelopmental Conditions | 7.2% | 7.5% | 10.6% | 6.0% | 0.01 | **0.12** | 0.05 |
| Obesity | 20.9% | 21.2% | 19.7% | 22.3% | 0.01 | 0.03 | 0.04 |
| Pneumonia | 5.0% | 4.6% | 6.7% | 3.8% | 0.02 | 0.07 | 0.06 |
| Hospitalized Stroke | 0.6% | 0.6% | 0.8% | 0.5% | 0.01 | 0.02 | 0.02 |
| **Prior COVID-19 Diagnosis** | | | | | | | |
| Any | 3.3% | 4.6% | 7.4% | 4.6% | 0.06 | **0.18** | 0.07 |
| Inpatient | 0.9% | 0.9% | 1.7% | 1.0% | 0.00 | 0.07 | 0.01 |
| Outpatient or Professional | 3.3% | 4.5% | 7.4% | 4.6% | 0.07 | **0.18** | 0.07 |

Abbreviations: FFS, fee-for-service; SMD, standardized mean difference; AMI, acute myocardial infarction; COPD, chronic obstructive pulmonary disease; COVID-19, coronavirus disease 2019

[a] Individuals included in this table were required to have 365 days of continuous enrollment prior to vaccination date in order to accurately capture medical history

[b] Characteristics of the general Medicare FFS population aged 65 years and older were assessed as of 12/10/2020

[c] Characteristics of the vaccination doses (e.g., Moderna vaccine doses) were assessed at the administration date of the first dose of each vaccinated individual

**Table S8. Characteristics of Pfizer-BioNTech and Moderna Vaccine Doses[a] Administered among Adults Aged 65 years and Older in Medicare[b], Dec 11, 2020 to Feb 27, 2021**

| Patient Characteristic | General 65+ FFS[c] | Moderna[d] | Pfizer | SMD - Comparison with General FFS | |
|---|---|---|---|---|---|
| | | | | Moderna | Pfizer |
| **Total** | **25,390,578** | **4,224,008** | **4,464,086** | | |
| **Nursing Home Residential Status** | | | | | |
| Nursing Home Resident | 2.3% | 4.4% | 11.9% | **0.12** | **0.38** |
| Non-Nursing Home Resident | 97.7% | 95.6% | 88.1% | **0.12** | **0.38** |
| **Age (years)** | | | | | |
| 65-74 | 53.8% | 46.3% | 38.2% | **0.15** | **0.32** |
| 75-84 | 32.6% | 37.1% | 38.1% | 0.09 | **0.11** |
| 85+ | 13.5% | 16.6% | 23.7% | 0.08 | **0.26** |
| **Sex** | | | | | |
| Female | 55.9% | 57.8% | 60.6% | 0.04 | 0.09 |
| Male | 44.1% | 42.2% | 39.4% | 0.04 | 0.09 |
| **Race/Ethnicity** | | | | | |
| Asian | 2.1% | 1.8% | 1.7% | 0.02 | 0.03 |
| Black | 6.8% | 3.9% | 4.7% | **0.13** | 0.09 |
| Hispanic | 1.6% | 0.7% | 0.7% | 0.08 | 0.08 |
| Alaskan Native/Native American | 0.5% | 0.7% | 0.7% | 0.03 | 0.04 |
| White | 85.0% | 88.7% | 88.4% | **0.11** | 0.10 |
| Other | 1.8% | 1.8% | 1.7% | 0.01 | 0.00 |
| Missing/Unknown | 2.3% | 2.4% | 2.0% | 0.01 | 0.02 |
| **Urban/Rural** | | | | | |
| Urban | 76.5% | 75.2% | 82.7% | 0.03 | **0.15** |
| Rural | 22.9% | 24.3% | 16.9% | 0.03 | **0.15** |
| Missing/Unknown | 0.6% | 0.5% | 0.4% | 0.02 | 0.03 |
| **Medical Conditions (0-365 Days Prior to Index Date)** | | | | | |
| Atrial Fibrillation | 13.7% | 15.8% | 18.5% | 0.06 | **0.13** |
| AMI Hospitalization | 0.7% | 0.6% | 0.8% | 0.01 | 0.01 |
| Bronchiectasis | 0.9% | 1.1% | 1.3% | 0.02 | 0.03 |
| Charlson Comorbidity Index >0 | 63.3% | 67.3% | 72.4% | 0.08 | **0.20** |
| Asthma w/o COPD | 4.8% | 5.2% | 5.0% | 0.02 | 0.01 |
| COPD | 12.0% | 11.7% | 13.3% | 0.01 | 0.04 |
| Coronary Revascularization | 0.9% | 0.9% | 0.8% | 0.00 | 0.01 |
| Diabetes | 27.0% | 26.9% | 28.1% | 0.00 | 0.03 |

| Patient Characteristic | General 65+ FFS[c] | Moderna[d] | Pfizer | SMD - Comparison with General FFS | |
|---|---|---|---|---|---|
| | | | | Moderna | Pfizer |
| Depression | 17.0% | 18.4% | 24.6% | 0.04 | **0.19** |
| Gout | 5.4% | 5.8% | 5.9% | 0.02 | 0.02 |
| Hospitalization | 13.1% | 12.8% | 17.4% | 0.01 | **0.12** |
| Hypertension | 67.8% | 72.2% | 74.8% | 0.10 | **0.15** |
| Interstitial Lung Disease | 1.9% | 2.1% | 2.3% | 0.02 | 0.03 |
| Impaired Mobility | 0.4% | 0.5% | 1.0% | 0.00 | 0.06 |
| Neurological/Neurodevelopmental Conditions | 7.2% | 8.0% | 11.6% | 0.03 | **0.15** |
| Obesity | 20.9% | 20.7% | 19.1% | 0.00 | 0.04 |
| Pneumonia | 5.0% | 4.9% | 7.4% | 0.01 | 0.10 |
| Hospitalized Stroke | 0.6% | 0.6% | 0.9% | 0.00 | 0.03 |
| **Prior COVID-19 Diagnosis** | | | | | |
| Any | 3.3% | 4.8% | 8.2% | 0.08 | **0.21** |
| Inpatient | 0.9% | 1.0% | 1.8% | 0.01 | 0.08 |
| Outpatient or Professional | 3.3% | 4.8% | 8.2% | 0.08 | **0.21** |

Abbreviations: FFS, fee-for-service; SMD, standardized mean difference; AMI, acute myocardial infarction; COPD, chronic obstructive pulmonary disease; COVID-19, coronavirus disease 20

[a] Janssen is not included because only 88 Janssen vaccine doses had been observed by February 27, 2021

[b] Individuals included in this table were required to have 365 days of continuous enrollment prior to vaccination date in order to accurately capture medical history

[c] Characteristics of the general Medicare FFS population aged 65 years and older were assessed as of 12/10/2020

[d] Characteristics of the vaccination doses (e.g., Moderna vaccine doses) were assessed at the administration date of the first dose of each vaccinated individual

**Table S9. Summary of Data Quality Checks for Acute Myocardial Infarction, Pulmonary Embolism, Disseminated Intravascular Coagulation, and Immune Thrombocytopenia identified via Diagnosis Codes after Pfizer-BioNTech vaccination in Medicare Shared Systems Database**

| Evaluations | Acute Myocardial Infarction (AMI) | Pulmonary Embolism (PE) | Disseminated Intravascular Coagulation (DIC) | Immune Thrombocytopenia (ITP) |
|---|---|---|---|---|
| Duplication of vaccines/outcomes | None | None | None | None |
| Unusual variability in claims accrual | None | None | None | None |
| Number of individuals with other outcomes (i.e., overlap between outcome populations) | 3.5% with PE<br>0.4% with DIC<br>0.2% with ITP | 4.9% with AMI<br>0.2% with DIC<br>0.1% with ITP | 15.4% with AMI<br>6% with PE<br>1.1% with ITP | 1.6% with ITP<br>0.3% with DIC<br>0.7% with PE |
| Changes in diagnosis criteria or guidelines in detecting outcomes or vaccines | None | None | None | None |
| Assess changes in payment policy of claims submission | None | None | None | None |

**Table S10. Summary of Primary and Sensitivity Rapid Cycle Analyses for Acute Myocardial Infarction, Pulmonary Embolism, Disseminated Intravascular Coagulation, and Immune Thrombocytopenia identified after Pfizer-BioNTech vaccination in Medicare Shared Systems Database**

| Analysis | Data Through Signal Date of Primary Analysis | | Data Through May 1, 2021 | | Statistical Signal | Last LLR / Critical Value |
|---|---|---|---|---|---|---|
| | Observed Cases | RR | Observed Cases | RR | | |
| **AMI** | | | | | | |
| Primary | 1526 | 1.42 | 6224 | 1.13 | Yes | 11.42/3.49 |
| Variation in monthly rates | 1526 | 1.24 | 6224 | 1.02 | No | 0.00/4.10 |
| Influenza-Vaccinated | 1526 | 1.41 | 6224 | 1.14 | Yes | 10.93/3.50 |
| **PE** | | | | | | |
| Primary | 1197 | 1.54 | 4538 | 1.36 | Yes | 23.76/3.77 |
| Variation in monthly rates | 1197 | 1.35 | 4538 | 1.25 | Yes | 14.15/4.03 |
| Influenza-Vaccinated | 1197 | 1.48 | 4538 | 1.30 | Yes | 15.65/3.73 |
| Peri-COVID | 1197 | 1.22 | 4538 | 1.10 | No | 0.00/3.59 |
| Inpatient-Only | 498 | 2.17 | 1876 | 1.45 | Yes | 63.23/4.90 |
| **DIC** | | | | | | |
| Primary | 82 | 1.91 | 181 | 1.30 | Yes | 6.45/6.03 |
| Variation in monthly rates | 82 | 1.59 | 181 | 1.16 | No | 0.00/5.05 |
| Influenza-Vaccinated | 82 | 1.87 | 181 | 1.30 | No | 0.14/5.15 |
| Peri-COVID | 82 | 1.54 | 181 | 1.06 | No | 0.00/4.98 |
| **ITP** | | | | | | |
| Primary | 644 | 1.44 | 691 | 1.42 | Yes | 5.87/5.07 |
| Variation in monthly rates | 644 | 1.37 | 691 | 1.35 | No | 2.15/5.07 |
| Influenza-Vaccinated | 644 | 1.28 | 691 | 1.26 | No | 0.03/5.17 |
| Peri-COVID | 644 | 1.64 | 691 | 1.62 | Yes | 6.86/5.77 |

Abbreviations: LLR, log likelihood ratio; RR, risk ratio
